# Hyper-Exponential Growth of COVID-19 during Resurgence of the Disease in Russia

**DOI:** 10.1101/2020.10.26.20219626

**Authors:** Hemanta K. Baruah

**Affiliations:** Department of Mathematics, The Assam Royal Global University, Guwahati, Assam, India

**Keywords:** Pandemic, infectious disease, epidemiological modeling

## Abstract

In Russia, COVID-19 has currently been growing hyper-exponentially. This type of a spread pattern was not seen during the first wave of the pandemic the world over. Indeed when the disease had first appeared, in the accelerating stage the spread pattern was observed to have followed a highly nonlinear pattern that could be said to be approximately exponential or sub-exponential. As to why in the resurgence the growth has become hyper-exponential is another matter. But this has been happening in Europe and how long this would continue cannot be predicted. It may so happen that in the countries in which retardation has already been taking place, there may be resurgence of the disease. It was observed that in the World as a whole, retardation was on the threshold during the second half of September. But if the resurgence happens to follow the hyper-exponential growth pattern in different countries, there may be resurgence in the World as a whole.

## Introduction

When COVID-19 had first appeared, it was observed that in the accelerating stage the disease had been spreading following a highly nonlinear pattern which was approximately exponential which might be termed as sub-exponential. In Europe, the spread of the disease almost came to a halt months ago. However, in Europe it has reappeared and this time unlike in the first wave the growth rate has been much faster than what was observed earlier. At the sub-exponential stage, forecasting about retardation was not difficult. In this article, we are going to show that the growth pattern in Russia is hyper-exponential, and it is not going to be easy to forecast when the growth would start retarding. Regarding hyper-exponential pattern we would now like to discuss a few things in short.

The Malthusian model [1] was the initial work on population dynamics in which it was stated that a population when unchecked grows exponentially following the pattern

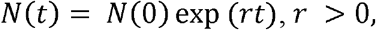

where *N*(*t*) is the population at time *t* and *r* is the exponential rate of growth of the population. This can equivalently be written as

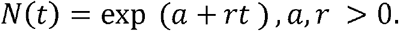

It was assumed in the model above that *r* is a constant. In other words, it was presumed that the rate of change of the natural logarithm of *N*(*t*) is a constant.

Fisher’s theorem on natural selection [2] states that *r* is a function of time *t* and

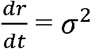

where *σ*^2^ was termed as the genetic variance in fitness, following

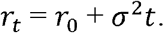

This finally gives

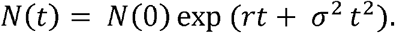

This in turn can be equivalently written as

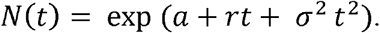

This is what was referred to hyper-exponential growth of a population. Fisher’s theorem was indeed the limit theorem of population dynamics which can be seen as a generalization of the Malthusian model. In Fisher’s model it is inherently presumed that the reason behind such a time dependent positive rate of growth is caused by genetic variations causing an additive effect with reference to time. Therefore in Fisher’s model, the rate of change of the natural logarithm of *N*(*t*) is of first degree in *t*. Accordingly, the population growth in this model is much faster than the exponential growth model due to Malthus.

It may be noted that all biological populations grow according to exponential law, whereas the human population grows following the hyper-exponential law which essentially moves ahead of the exponential one. A hyper-exponential equation that allows describing dynamics of human population was in fact obtained in [3].

In this article, we are going to demonstrate that in Russia currently the rate of growth instead of decreasing with time is actually increasing. This would show that currently in Russia the growth is hyper-exponential.

The classical epidemiological models [4, 5, 6] assume exponential growth in the accelerating stage of the spread of an epidemic. Indeed, in the accelerating stage an exponential growth pattern should be only theoretically possible, for if the increase is actually exponential the epidemic would never come to a halt. Therefore, an epidemic growth can only be approximately exponential or sub-exponential with the rate of growth decreasing in time. This decreasing rate of growth would be reflected in the estimated values of the growth parameter *r* with respect to time.

In this study, we shall use data from Worldometers.info [7]. We would like to mention here that the data in this source get edited after publication, which results in minor changes in the data.

Therefore there may be seen some minor changes in the data that we are going to tabulate in this article.

## Methodology

We are going to demonstrate that the current growth rate of COVID-19 in Russia is following the hyper-exponential law. For that we would define

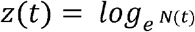

where *N*(*t*) is the cumulative total number of COVID-19 cases in Russia at time *t*. We shall first assume that the spread is approximately following the function

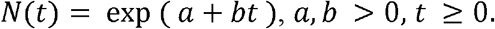

Indeed for *N*(*t*) strictly exponential following the pattern shown above

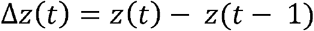

would be constant.

But in the case of an epidemic, the function *N*(*t*) can be only approximately exponential. Once the cumulative total number of cases of the epidemic enters into the nearly exponential phase, Δ*z*(*t*) starts to follow a reducing trend. It was observed in [8, 9, 10, 11, 12] that when the pandemic continues to grow nearly exponentially Δ*z*(*t*) continues to decrease in time. In [13] it was shown that in six European countries in general and in Russia in particular, the values of Δ*z*(*t*) were not showing decreasing trend. It was shown empirically that in Russia the situation was most serious. In this article, we shall now find the linear regression equation of Δ*z*(*t*) on time *t*. Based on the regression equation, after testing statistically we would infer whether the COVID-19 growth pattern in Russia is exponential or hyper-exponential.

### Analysis

In what follows we are going to see first whether the values of Δ*z*(*t*) are following a decreasing trend, or an increasing trend, or near constancy. As mentioned earlier, during the sub-exponential stage of growth of COVID-19, Δ*z*(*t*) was observed to have followed a decreasing trend. It was shown in [11] that from June 23 to August 21, in India Δ*z*(*t*) had followed the following three linear equations established using the method of least squares. The study was made for 60 days, separated into three equal parts of 20 days each. From June 23 to July 12, the equation was

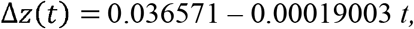

from July 13 to August 1, it was

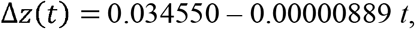

and from August 2 to August 21, it was

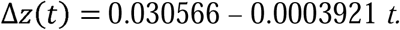

Statistical validity of all three equations was tested. It was statistically concluded that during the first 20 days the coefficient of regression of Δ*z*(*t*) on *t* was negative, during the next 20 days it was insignificant and in the last 20 days the regression coefficient was negative again. Using the equation fitted from data from August 2 to August 21, a forecast was made in [11] that starting from August 2, within no more than 78 days retardation would start in India. That forecast could be seen to be true.

In Table-1, we have tabulated the values of *N*(*t*), *Z*(*t*) and Δ*z*(*t*) for Russia for the period of 20 days from 2 October to 21 October. In Fig.1 we have depicted the observed and the expected values of Δ*z*(*t*).

**Table 1:** Values of Δ*z*(*t*) in Russia from 2 October to 21 October

| Date | $N(t)$ | $z(t)$ | $\Delta z(t)$ |
| --- | --- | --- | --- |
| 1-Oct | 1185231 | 13.98545 | ---- |
| 2-Oct | 1194643 | 13.99336 | 0.00791 |
| 3-Oct | 1204502 | 14.00158 | 0.008219 |
| 4-Oct | 1215001 | 14.01026 | 0.008679 |
| 5-Oct | 1225889 | 14.01918 | 0.008921 |
| 6-Oct | 1237504 | 14.02861 | 0.00943 |
| 7-Oct | 1248619 | 14.03755 | 0.008942 |
| 8-Oct | 1260112 | 14.04671 | 0.009162 |
| 9-Oct | 1272238 | 14.05629 | 0.009577 |
| 10-Oct | 1285084 | 14.06633 | 0.010047 |
| 11-Oct | 1298718 | 14.07689 | 0.010554 |
| 12-Oct | 1312310 | 14.0873 | 0.010411 |
| 13-Oct | 1326178 | 14.09781 | 0.010512 |
| 14-Oct | 1340409 | 14.10849 | 0.010674 |
| 15-Oct | 1354163 | 14.11869 | 0.010209 |
| 16-Oct | 1369313 | 14.12982 | 0.011126 |
| 17-Oct | 1384235 | 14.14066 | 0.010838 |
| 18-Oct | 1399334 | 14.15151 | 0.010849 |
| 19-Oct | 1415316 | 14.16286 | 0.011356 |
| 20-Oct | 1431635 | 14.17433 | 0.011464 |
| 21-Oct | 1447335 | 14.18523 | 0.010907 |

It is apparent that the values of Δ*z*(*t*) are showing an increasing trend. We shall therefore move ahead to check whether this increasing trend is linear. The linear regression equation of Δ*z*(*t*) for Russia for the period from 2 October to 21 October has been found to be

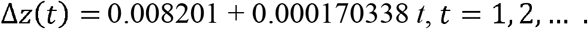

It is apparent that the linear regression coefficient of Δ*z*(*t*) on *t* is positive, and this is evidence that the growth pattern of C0VID-19 in Russia during the period from 2 October to 21 October is perhaps hyper-exponential. We now proceed to see whether this equation is statistically acceptable or not. We have done the statistical tests of significance of the null hypothesis *H*_0_ : *ρ*= 0 against the two sided alternative hypothesis *H*_1_ : *ρ* ≠ 0 where *ρ* is the population correlation coefficient between the variables Δ*z*(*t*) and *t*. We have found that the Error Sum of Squares = 2.21347E-06 and the Total Sum of Squares = 2.15084E-05. The ratio gives us (1 − *r*^2^) = 0.102911, and hence *r*^2^= 0.897089. Using the Student’s *t* test, with

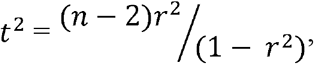

where *n* = 20, we have found that the calculated value of *t* is equal to 12.526309 which is far greater than the two sided theoretical value of *t* (= 2.10092), at 5% probability level of significance for 18 degrees of freedom. Therefore we conclude that the null hypothesis is to be rejected, and that there is a significant linear relationship between Δ*z*(*t*) and *t*. This is equivalent to concluding that the value of the regression coefficient (= + 0.000170338) is significantly different from 0.0. Indeed we would like to mention at this point that the theoretical value of *t* for 18 degrees of freedom at 1% probability level of significance is 2.87844, and at 0.1% probability level of significance it is 3.9651. This means that we can be 99.9% sure that the population correlation coefficient is very highly significantly different from zero.

In Table-2 we have shown the forecasts of the cumulative total number of cases for a short period from 22 October to 5 November. We have depicted the forecasts in Fig. 2.

**Table 2:** Forecasts for Russia from 22 October to 5 November

| Date | $\Delta z(t)$ | $z(t)$ | $N(t)$ |
| --- | --- | --- | --- |
| 22-Oct | 0.011778 | 14.19701 | 1464482 |
| 23-Oct | 0.011948 | 14.20896 | 1482085 |
| 24-Oct | 0.012119 | 14.22108 | 1500156 |
| 25-Oct | 0.012289 | 14.23337 | 1518705 |
| 26-Oct | 0.012459 | 14.24583 | 1537745 |
| 27-Oct | 0.01263 | 14.25846 | 1557290 |
| 28-Oct | 0.0128 | 14.27126 | 1577351 |
| 29-Oct | 0.01297 | 14.28423 | 1597943 |
| 30-Oct | 0.013141 | 14.29737 | 1619080 |
| 31-Oct | 0.013311 | 14.31068 | 1640775 |
| 1-Nov | 0.013481 | 14.32416 | 1663044 |
| 2-Nov | 0.013652 | 14.33781 | 1685904 |
| 3-Nov | 0.013822 | 14.35163 | 1709368 |
| 4-Nov | 0.013992 | 14.36563 | 1733454 |
| 5-Nov | 0.014163 | 14.37979 | 1758180 |

**Fig. 1:**
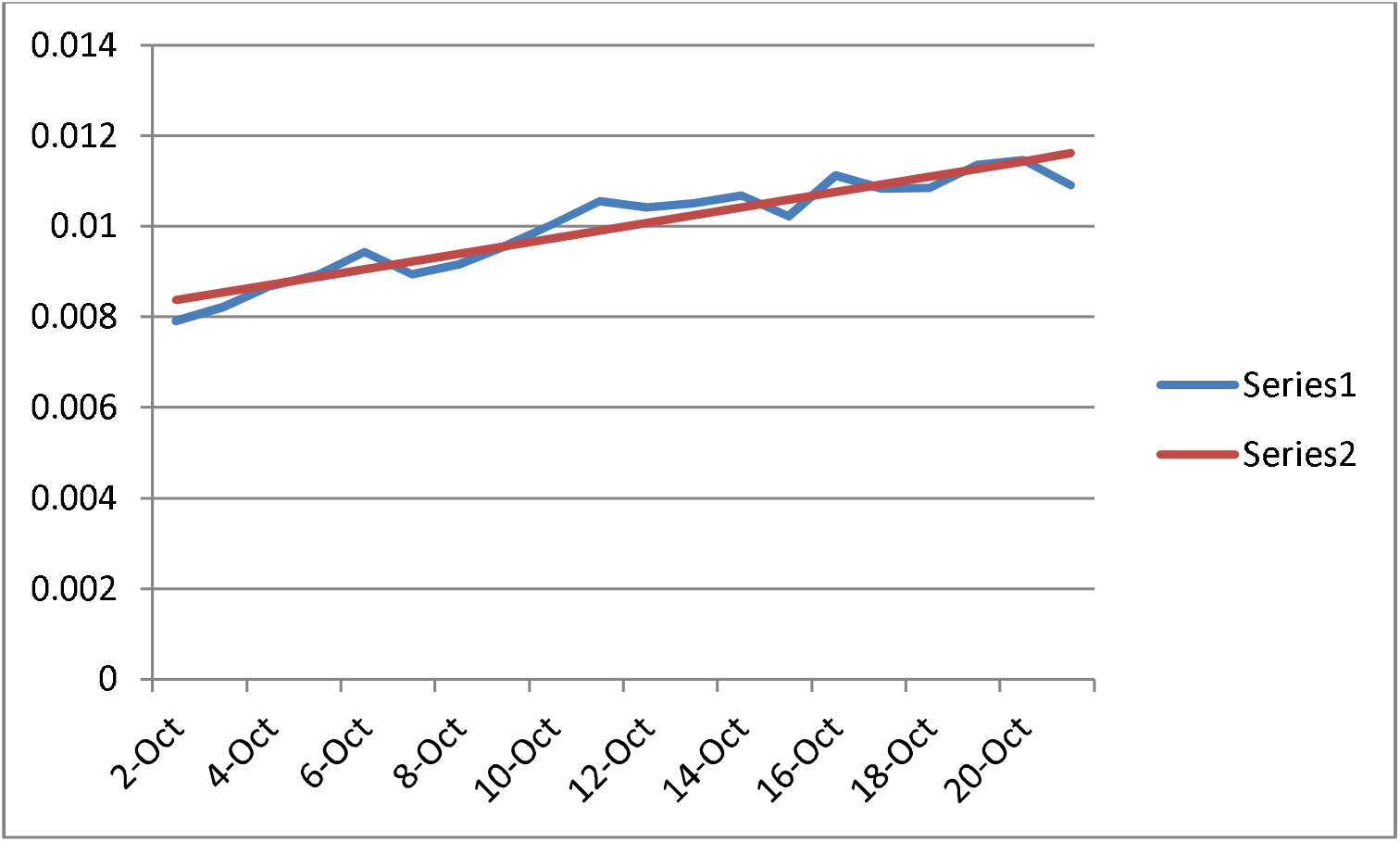
Observed and Expected Δ*z*(*t*) in Russia from 2 October to 21 October Series 1: Observed Values, Series 2: Expected values

**Fig. 2:**
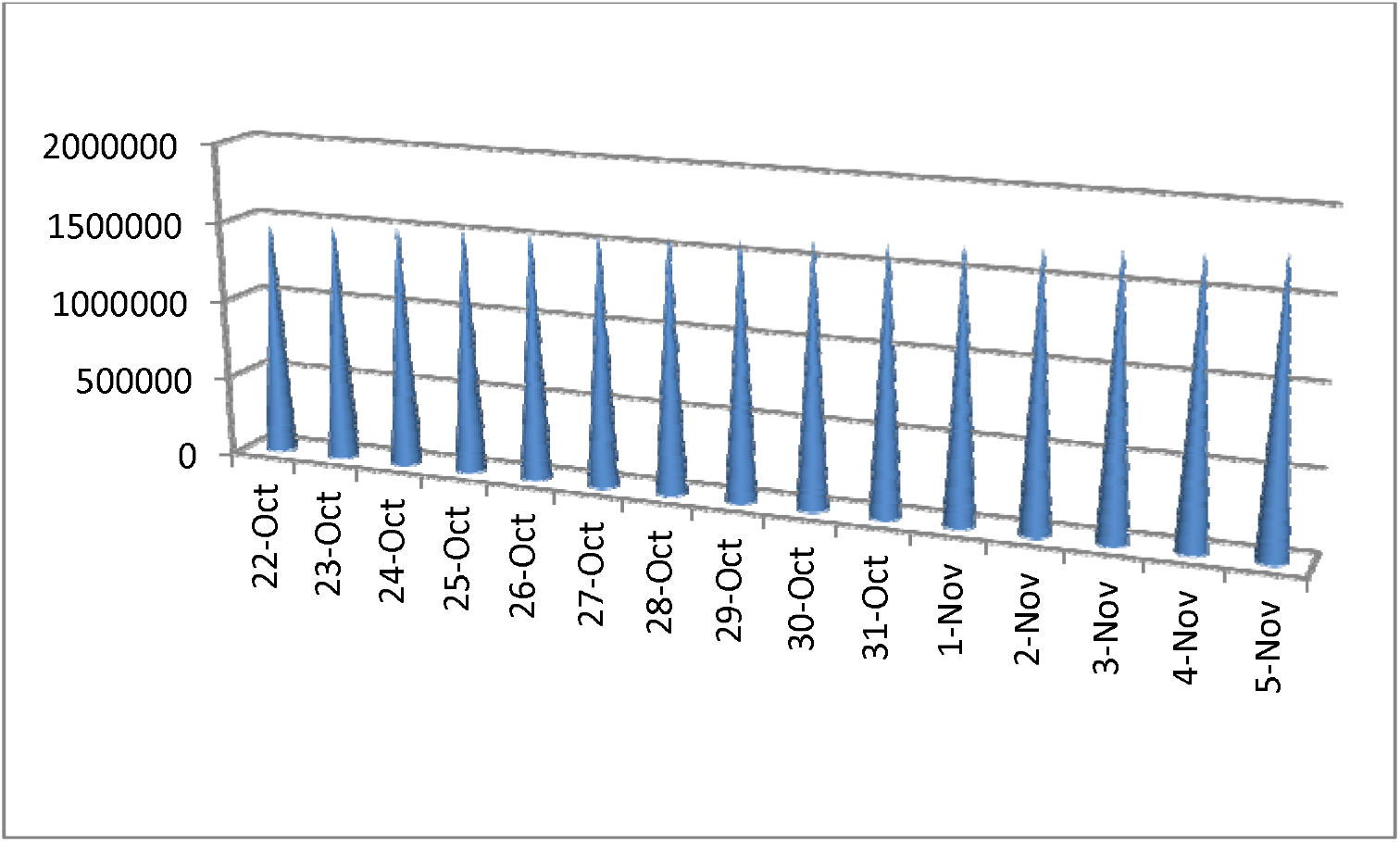
Forecasts of the Cumulative Total Number of Cases in Russia from 22 October to 5 November

It is clear that in this second wave, the COVID-19 growth in Russia is hyper-exponential. When approximately would the situation change towards betterment is difficult to say.

Thus we have demonstrated that Fisher’s population dynamics model is at work in Russia with reference to spread of COVID-19. Hyper-exponential growth is very unusual in this case because when the disease had first appeared it was following the Malthusian model of population dynamics which states that the growth is exponential.

## Conclusions

Whatever be the reason behind, the spread of COVID-19 in Russia is not following the pattern that it followed when it had first appeared in the World. During the accelerating stage, the growth was sub-exponential everywhere in the first wave, but in this second wave, the disease has started to grow much faster than that in the first wave. In the first wave, predictions could be made regarding around what time the retardation would start. But in the second wave the growth has been confirmed to be hyper-exponential. Therefore it is now difficult to say when the retardation would restart.

## Data Availability

The data have been taken from Worldometers.info.

https://www.worldometers.info

